# Lifestyles, left atrial structure and function, and cognitive decline in adults with the metabolic syndrome

**DOI:** 10.1101/2023.06.23.23291821

**Authors:** Ines Gonzalez Casanova, Ángel M. Alonso-Gómez, Dora Romaguera, Estefanía Toledo, Linzi Li, Elena Fortuny, Luis López, Raúl Ramallal, Jordi Salas-Salvadó, Lucas Tojal-Sierra, Olga Castañer, Alvaro Alonso

**Affiliations:** Department of Applied Health Science, Indiana University Bloomington School of Public Health, Bloomington, IN, USA; Bioaraba Health Research Institute; Osakidetza Basque Health Service, Araba University Hospital; University of the Basque Country UPV/EHU; Vitoria-Gasteiz, Spain; CIBER Consortium, M.P. Physiopathology of Obesity and Nutrition (CIBERObn), Carlos III Health Institute (ISCIII), Madrid, Spain; Health Research Institute of the Balearic Islands (IdISBa), Palma de Mallorca, Spain; Department of Preventive Medicine and Public Health, University of Navarra, Pamplona, Spain; IdiSNA, Navarra Institute for Health Research, Pamplona, Spain; Department of Epidemiology, Rollins School of Public Health, Emory University, Atlanta, GA, USA; Cardiology Service, Manacor Hospital, Palma de Mallorca, Spain; Cardiology Service, Son Espases University Hospital, Palma de Mallorca, Spain; Department of Cardiology, University Hospital of Navarra, Servicio Navarro de Salud Osasunbidea, Pamplona, Spain; Universitat Rovira i Virgili, Department of Biochemistry and Biotechnology, Human Nutrition Unit, Reus, Spain; Institut d’Investigació Sanitària Pere Virgili (IISPV), Human Nutrition Unit, Reus, Spain; Cardiovascular risk and nutrition research group, Institut Hospital del Mar d’Investigacions Mèdiques (IMIM), Barcelona, Spain

## Abstract

Evidence supports associations of lifestyle–including diet and physical activity—and weight with cognitive functioning, but the pathways responsible for these associations have not been fully elucidated. Because healthier lifestyles have been associated with better left atrial structure and function, which in turn is associated with better cognitive functioning, we tested the hypothesis that left atrial structure and function is a potential mediator of the association between lifestyles and cognition. We included 476 participants with overweight or obesity and metabolic syndrome from three centers in Spain who underwent lifestyle assessment and transthoracic echocardiography at baseline and had repeated measurements of the Trail Making A test, a measure of executive function, at baseline and at the two-year follow-up. We conducted mediation analyses to test if measures of left atrial structure and function mediated associations between adherence to the Mediterranean diet scores, physical activity, or weight at baseline, and two-year change in Trail Making A scores. The analysis did not find an effect between these factors and Trail Making A scores, and no indirect effects mediated through the echocardiographic measurements. The modest sample size in this analysis is a limitation, and larger studies should be conducted to determine potential cardiovascular factors mediating the association between lifestyle and cognition.

## Introduction

Cognitive impairment, defined as having difficulties with memory, learning or cognitive tasks beyond those expected based on age and educational level,^1^ has become a primary health concern for aging populations. It is estimated that approximately 11% of US adults over 65 years suffer from dementia and 23% have mild cognitive impairment.^2^ Elements of a healthy lifestyle including healthy eating, maintaining weight, and engaging in physical activity have been associated with less cognitive decline and dementia prevention primarily in observational studies.^3^ In terms of healthy eating, the strongest associations have been shown with adherence to the Mediterranean diet or Mediterranean-style dietary patterns like the MIND diet.^4^ Two meta-analyses of observational studies concluded that adherence to the Mediterranean diet was associated with prevention of cognitive impairment, dementia and Alzheimer’s disease.^5,6^ Similarly, obesity and physical inactivity are two out of 12 modifiable risk factors that account for 40% of worldwide dementias.^7^ Overweight has been associated with higher risk of mild cognitive decline and dementia,^8^ and regular moderate physical activity is associated with better cognitive function in adults older than 60 years.^9^ In fact, physical inactivity was identified as having the highest population attributable risk for Alzheimer’s disease in the US, Europe, and the United Kingdom.^10^

Healthy eating, weight and physical activity are also associated with healthier left atrial (LA) structure and function.^11^ In turn, markers of atrial remodeling and fibrosis, such as enlarged LA size and volume, and abnormal LA function have been associated with cognitive decline and dementia.^12^ In a recent analysis among a subsample of participants of the PREDIMED PLUS trial, we found that larger left atrial volume index, lower peak longitudinal strain, and higher stiffness index were associated with 2-year worsening performance in Trail-Making Test A, a measure of executive function.^13^ Hence, the objective of this analysis was to test the hypothesis that associations of weight and lifestyle factors with 2-year cognitive decline were mediated by measurements of LA structure and functioning in this same sample of participants with overweight or obesity.

## Methods

We included a subsample of participants form the PREDIMED PLUS trial, an ongoing multicenter randomized controlled trial aimed at preventing cardiovascular disease in overweight or obese adults with metabolic syndrome (ISRCTN89898870).^14^ For this analysis we included a sub-sample of 476 participants from three recruiting centers (University of Navarra, Araba University Hospital, Son Espases University Hospital) who underwent transthoracic echocardiography at baseline and who also had information on the Trail Making A test at baseline and at the two-year follow-up.

Trail Making Test measures processing speed and executive function.^15^ The Trail Making Test A, where participants are asked to connect numbers 1 to 25 in the correct order, is meant to assess cognitive processing skills. The score for this test is obtained based on the number of seconds that it takes to complete the test. Scores were multiplied by -1 for higher scores to represent better performance. Two-year follow up scores were standardized by subtracting the baseline mean and dividing them by the baseline standard deviations. To calculate 2-year cognitive change, standardized baseline scores were deducted from 2-year follow-up scores *([(2y score - baseline X)/baseline SD] - [(baseline score - baseline X)/baseline SD]*)

Echocardiographic examinations were performed at baseline using an ultrasound scanner Vivid 7 or Vivid 9 (General Electric Healthcare) following procedures described elsewhere.^16^ Adherence to the Mediterranean diet was assessed a 17-item short screener for adherence to the energy-reduced Mediterranean diet.^17^ Metabolic equivalents (METs) per day of moderate to vigorous physical activity were estimated using the short version of the Minnesota leisure time physical activity questionnaire.^18^ Height and weight were measured during the baseline visit. Participants’ sociodemographic and health characteristics were assessed at baseline via a face-to-face questionnaire.

We conducted mediation analyses using the methodology and SAS macro developed by Valeri and Vanderweele (2013).^19^ Three different models were conducted to estimate the total effects of adherence to the Mediterranean diet score, MVPA metabolic equivalents per day, and body mass index on Trail Making Test A, as well as to decompose the effect into direct effects and indirect effects mediated through echocardiographic variables that had been shown to be associated with change in cognitive scores in our previous analysis (left atrial volume index, peak systolic longitudinal strain, conduit strain, contractile strain, and stiffness index).^20^ Exposure variables were converted into z-scores by multiplying individual values times the mean and dividing it by the standard deviation. Hence, model estimates represent the two-year difference in Trail Making Test A standard deviations per a standard deviation change on the exposure variable. Models were adjusted for age, sex, site, number of people living in the household, employment status (yes or no), education (primary, high school, professional), smoking status (never, former, current), diagnosis of arrhythmia, self-reported diagnosis of diabetes, self-reported diagnosis of depression at baseline, and intervention group.

## Results

Participants were 61% male, had an average age of 65 years, and had on average 12 years of schooling (Table 1). The Trail Making Test A scores were 48.1 (21.2) at baseline and 49.9 (23.9) at the 2-year follow-up visit (difference: 1.8, 95%CI 0.0, 3.7). We did not find evidence for an effect of adherence to the Mediterranean diet, physical activity or body mass index on 2-year change in the Trail Making Test A (Table 2, Total effect column). Similarly, using effect decomposition, we did not find any significant direct effect or echocardiography-mediated indirect effect of adherence to the Mediterranean diet, physical activity, or body mass index on the outcome (Table 2).

**Table 1:** Sociodemographic and health characteristics of participants included in the analysis of echocardiographic markers of left atrial structure and change in Trail Making test A scores (n=476)

| **% or mean (SD)** | **Total (476)** |
| --- | --- |
| Site |  |
| Mallorca | 27.1 |
| Navarra | 20.8 |
| Vitoria | 52.1 |
| Age (years) | 65.2 (4.9) |
| Married | 77.9 |
| People living in the household | 1.3 (1.0) |
| Schooling (years) | 12.0 (5.2) |
| Employed | 17.7 |
| Health status |  |
| Body Mass Index (kg/m<sup>2</sup>) | 32.5 (3.3) |
| Diabetes (self-reported) | 22.7 |
| Hypertension (self-reported) | 83.6 |
| Depression (self-reported) | 17.9 |
| Arrhythmia (self-reported) | 5.5 |
| Health Behaviors |  |
| Current Smoker | 9.7 |
| Former Smoker | 51.3 |
| Mediterranean Diet Score (17-item screener) | 7.5 (2.9) |
| Moderate to vigorous physical activity (MET-min/day) | 269 (318) |
| General Echocardiographic Measures of Left Atrial Substrate |  |
| Volume Index (mL/m<sup>2</sup>) | 23.3 (7.5) |
| Peak Systolic Longitudinal Strain (%) | 27.3 (6.8) |
| Conduit Strain (%) | -11.9 (4.4) |
| Contractile Strain (%) | -15.4 (4.9) |
| Stiffness Index (U) | 0.4 (0.2) |
| Trail Making Test A (seconds) |  |
| Baseline | 48.1 (21.2) |
| 2-year difference | 1.9 (20.9) |

**Table 2:**
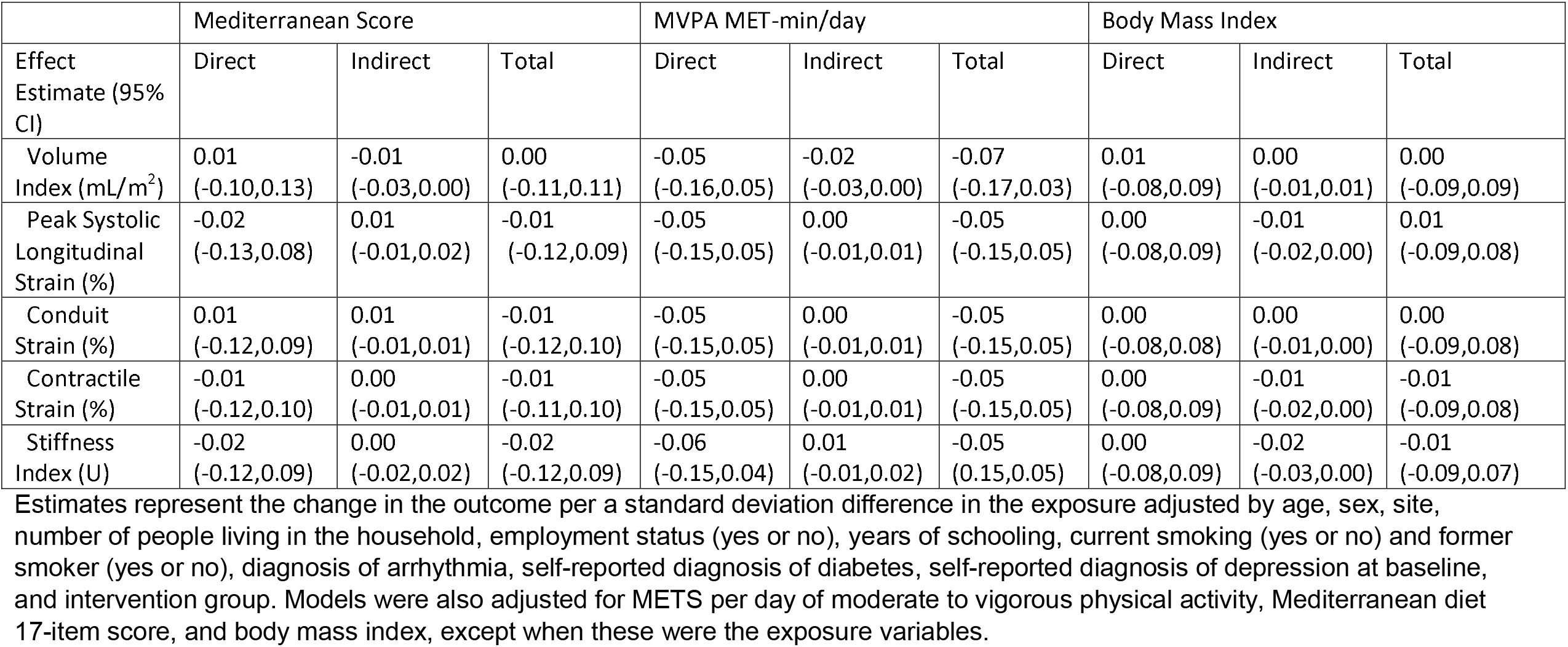
Analysis of left atrial volume and strain lifestyle variables as mediators in the association between (body mass index, Mediterranean diet score, METs in moderate to vigorous physical activity/day), and Trail Making Test 2-year change.

## Discussion

Based on previous evidence of the association between diet, physical activity, and body mass index with cognitive decline, and on our previous findings demonstrating associations between LA markers and cognitive decline in this cohort, we tested the hypothesis that LA markers mediated the association between these lifestyle factors and cognitive decline. Results from this analysis among 476 persons at high cardiovascular risk showed no association of adherence to the Mediterranean diet, daily moderate to vigorous physical activity, or body mass index with 2-year change in the Trail Making Test A. Consistently, there was no indirect association mediated by left atrial measurements. This result was unexpected, especially for adherence to the Mediterranean diet, which has been shown in several studies to be associated with cognitive function. ^5,6^ Potential explanations are the limited sample size, a short follow-up of only two years leading to imprecise estimates of effect, and that associations tested in this analysis were specific to 2-year change in a measure of executive function, while previous studies have looked at overall scores, having mostly used the Mini-mental Examination that measures overall cognitive functioning.^5,6^ Another difference with previous studies was that this analysis was limited to older adults with the metabolic syndrome.

Strengths of this analysis include the good retention of study participants, repeated measurements of cognition, information available on several sociodemographic and health covariates, and the use of echocardiography to assess LA structure and function. Potential limitations include the short follow-up time of only 2-years, the lack of precision in the estimates due to the modest sample size, and the exclusion of participants with cardiovascular disease or low cognitive functioning based on the design of the trial. Other analysis that include longer follow-ups, larger samples, based on randomized designs, and including diverse measurements of cognitive functioning are needed to elucidate the potential mediating role of LA measurements in the associations between lifestyle and cognitive functioning.

## Data Availability

This is a secondary analysis of the PREDIMED PLUS trial, which is ongoing. Data will be available as described on the Data Management Plan and Data Sharing Policy (link below).

https://www.predimedplus.com/wp-content/uploads/2018/10/PREDIMED_PLUS_V1_6_Data_26_4_18_ManagPlan_and_data_sharing_Policy.pdf

## Acknowledgements

The authors especially thank the PREDIMED-Plus participants for the enthusiastic collaboration, the PREDIMED-Plus personnel for outstanding support, and the personnel of all associated primary care centers for the exceptional effort. CIBEROBN, CIBERESP, and CIBERDEM are initiatives of Instituto de Salud Carlos III (ISCIII), Madrid, Spain. The authors also thank the PREDIMED-Plus Biobank Network as a part of the National Biobank Platform of the ISCIII for storing and managing the PREDIMED-Plus biological samples.

